# Predicting the *When*: Multimodal AI for Time-to-Recurrence Analysis After Atrial Fibrillation Ablation

**DOI:** 10.64898/2026.05.12.26353055

**Authors:** Minglang Yin, Changxin Lai, Ritu Yadav, Jenna A. Milstein, Linh Thi My Tran, Calvin O’Donnell, Sarah Schumacher, Craig Cronin, Robert Weinstein, Carolyna Yamamoto Alves Pinto, Zan Ahmad, Shiyi Chen, Arthur Lefebvre, Jooyoung Ryu, Audrey Lacy, Alana Thi Yee, Jiwoo Noh, Eugene Kholmovski, Mauro Maggioni, Hugh Calkins, David Spragg, Natalia Trayanova

## Abstract

**Background:** Catheter ablation is the most effective rhythm control strategy for atrial fibrillation (AF); however, recurrence remains common. Current post-ablation management follows largely population-level protocols, constrained by the absence of tools that can anticipate not merely *whether*, but *when*, an individual patient will experience recurrence. The emergence of multimodal artificial intelligence (AI) presents a new opportunity to address this unmet clinical need.

**Objective:** To develop a predictive model for time-to-AF-recurrence post-ablation using pre-procedural bi-atrial imaging, clinical covariates, and procedural characteristics, within a novel multimodal AI and survival analysis framework.

**Methods:** We analyzed a retrospective cohort of 437 AF patients who underwent catheter ablation with follow-up censored at 36 months. **MARTA-AF** *(Multimodal AI Recurrence and Time-to-event Analysis post-Ablation in AF)* was trained on pre-procedural bi-atrial images, and covariates/procedural characteristics, and integrated into a survival model to generate time-varying recurrence probability estimates. Model interpretability was achieved by quantifying contribution of covariates/procedural characteristics to predicted survival probabilities.

**Results:** MARTA-AF successfully predicted time-varying recurrence risk up to three years post-ablation. Patients were effectively stratified into low– and high-risk groups, with statistically significant discrimination sustained over the follow-up period. The model demonstrated consistent performance across clinically relevant subgroups, including sex, age, and AF type. Incorporation of right atrial shape features improved time-to-AF-recurrence prediction. Interpretability analyses identified key recurrence predictors.

**Conclusions:** MARTA-AF delivers individualized, time-varying AF recurrence risk forecasts and enables stratification into clinically meaningful risk groups. This framework has the potential to transform post-ablation management into a proactive paradigm and to support informed clinical decision-making prior to ablation.

## Introduction

Catheter ablation is the most effective rhythm control strategy for atrial fibrillation (AF)^1^, yet recurrence rates remain substantial, ranging from 20% to 50% depending on AF type and patient substrate^2,3^. Current post-ablation management follows largely population-level protocols^4^ — empirical anticoagulation continuation, fixed-duration antiarrhythmic therapy, and arbitrary surveillance intervals — because clinicians lack the tools to anticipate *when*, not merely *whether*, an individual patient will recur. This distinction is clinically consequential.

Predicting time-to-recurrence, rather than binary AF recurrence alone, enables a fundamentally different paradigm: prospective, patient-specific decision-making anchored to a predicted clinical timeline. Anticoagulation management stands as perhaps the most immediately impactful application. Many patients remain on anticoagulants indefinitely post-ablation due to uncertainty about durable sinus rhythm^5^, accepting chronic bleeding risk as the cost of stroke prevention^4^. A reliable time-to-recurrence estimate could provide evidence-based justification for discontinuation in patients predicted to maintain sinus rhythm durably, while ensuring that patients predicted to recur early never enter a vulnerable anticoagulation gap — the precise window in which stroke risk is highest.

Beyond anticoagulation, predicted AF recurrence timing reframes several other management decisions. The standard 90-day blanking period is a biological approximation; knowing whether a patient is likely to recur within or well beyond that window transforms how early post-ablation arrhythmias are interpreted and acted upon. Similarly, repeat AF ablation is currently almost exclusively reactive, performed after symptomatic relapse and often after progressive atrial remodeling has already compounded the substrate. Time-to-recurrence prediction could define a patient-specific optimal re-intervention window — early enough to prevent remodeling, late enough for lesion maturation — converting redo procedures from a reactive rescue to a planned, strategically timed intervention. Antiarrhythmic drug duration and the intensity of remote monitoring could likewise be calibrated to each patient’s predicted trajectory, concentrating resources in high-risk windows rather than applying them uniformly across a heterogeneous population.

Collectively, these applications represent a shift from reactive to anticipatory post-ablation care. An AI model capable of predicting post-ablation time-to-AF-recurrence from pre-procedural imaging and clinical data would not simply add prognostic information — it would provide the actionable timeline that clinicians currently lack, enabling individualized, scheduled management decisions that population-level protocols cannot support. AI has been previously used only to predict *whether* AF will recur post-ablation within a fixed time period, such as one-year post-ablation leveraging a combination of medical data, such as clinical covariates, CT images^6,7^, ECG signals^8^, or intracardiac electrogram^9^. Yet, a predictive AI that predicts *when* the AF recurrence is likely to occur post-procedure is currently lacking.

The goal of this study is to develop a predictive model for time-to-AF-recurrence post-ablation using pre-procedural bi-atrial imaging and clinical data, and procedural characteristics, within a novel multimodal AI and survival analysis framework. A distinguishing aspect of this framework is the inclusion of the right atrium (RA) alongside the left atrium (LA) as imaging inputs — a departure from conventional practice. While the RA has been implicated in AF recurrence^10,11^, image acquisition and ablation strategies have historically centered on the LA. Both clinical risk indices and prior AI models have relied exclusively on LA geometric features; the incorporation of RA imaging therefore represents a distinct methodological advance. We term this combined multimodal AI and survival analysis framework **MARTA-AF** *(Multimodal AI Recurrence and Time-to-event Analysis post-Ablation in AF)*. **MARTA-AF** generates time-to-recurrence risk forecasts up to three years post-ablation. Model performance is benchmarked against established risk scores and evaluated across multiple patient populations. Using **MARTA-AF**-derived risk estimates, patients are stratified into high– and low-risk groups and their clinical characteristics examined. Finally, an AI interpretability analysis quantifies the contribution of individual covariates/procedural characteristics (or referred to as “tabular data”) to predicted survival probabilities, enhancing transparency and supporting clinical trustworthiness.

## Methods

### Study Population

The target population consisted of AF patients who underwent catheter ablation. A total of 437 patients were collected in the Johns Hopkins Hospital (JHH) AF ablation registry. Our retrospective cohort included patients who underwent both first-time (de novo) and repeat ablations. Patients were excluded if they met any of the following criteria: absence of post-procedural follow-up, missing or incomplete CT scans, images with filling defects, moderate to severe mitral regurgitation, incomplete procedures, or patients who had received the left atrial appendage occlusion procedure. All patients received pulmonary vein isolation (PVI); additional non-pulmonary vein targets (linear lesions; low voltage areas) were ablated at the operator’s discretion.

### Input Data

A schematic of the image processing pipeline is presented in the left panel of Figure 1. Contrast-enhanced CT scans were first acquired from several commercially available scanners, including Toshiba Aquilion Precision, Toshiba Aquilion ONE, Siemens Healthineers syngo, and Siemens Definition at JHH. Then, the left and right atrium (RA) blood pool was segmented by our open-source AI algorithm^12^, followed by manual correction by experienced operators. The segmented images were then interpolated from raw CT resolutions onto a template with pixel dimensions of 1.5 mm × 1.5 mm × 2.2 mm. For network training, post-processed images were binarized, with the bi-atrial blood pool labeled as 1 and the surrounding tissue labeled as 0.

**Figure 1:**
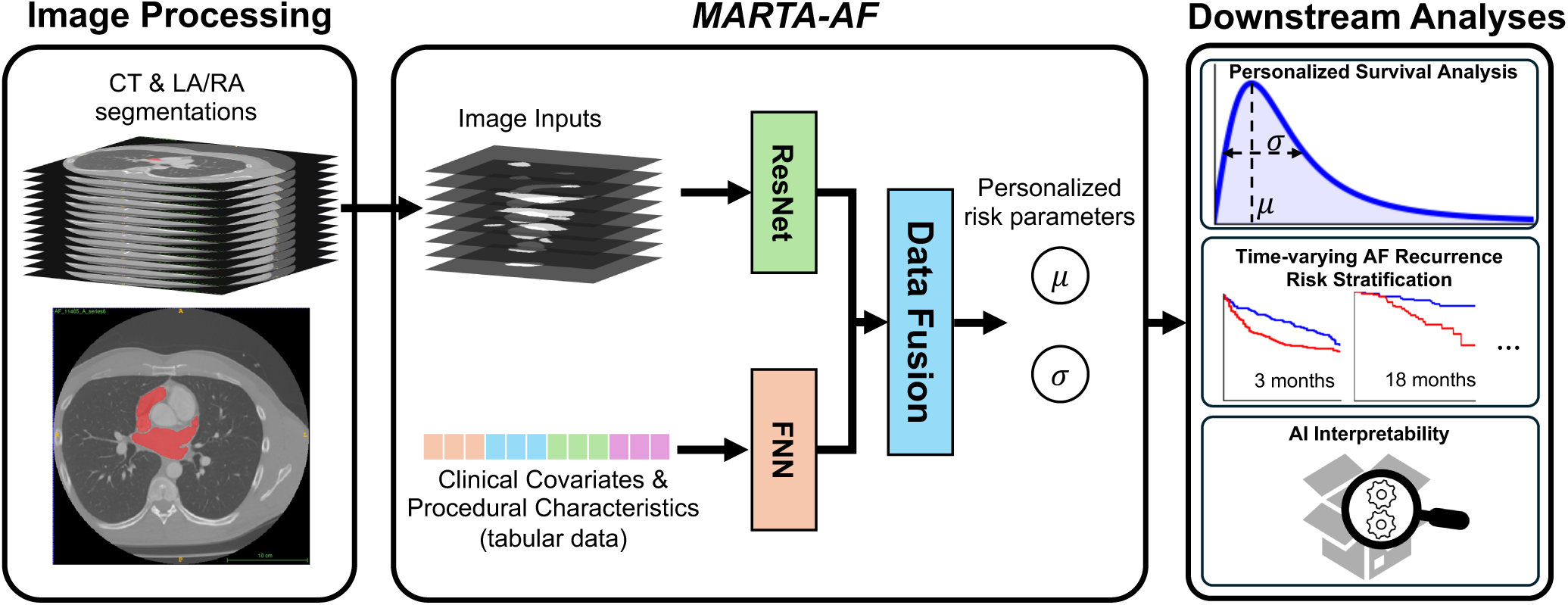
Schematic of image processing, MARTA-AF and its downstream analyses for time-to-recurrence analysis of AF. Left: Contrast-enhanced CT scans in AF patients were collected, followed by segmentation of the left and the right atrium blood pool. Middle: MARTA-AF and its input data. MARTA-AF utilized a feedforward neural network (FNN, red) and a ResNet to analyze tabular data (clinical covariates and procedural characteristics) and CT segmentation, and a FNN-based data fusion module (blue) to predict personalized risk parameters (μ and σ) by integrating multimodal information. Right: Downstream analyses were performed, including personalized survival analysis of AF recurrence using the predicted μ and σ, patient stratification at multiple time points based on their recurrence risk, and AI interpretability assessment.

In addition to medical images, the AI model took a list of clinical covariates as input (Figure 1, middle panel), including age, sex, height, weight, BMI, stroke, coronary artery diseases (CAD), OSA, hypertension, diabetes, AF type (persistent or paroxysmal) AF duration in years, left atrial diameters (LADs), pre-procedural left ventricular ejection fraction (LVEF), and *CHA*_2_*DS*_2_ − *VASc* score, as well as ablation procedural characteristics, such as ablation lesion (PVI only or with more lesion), catheter type (radiofrequency or others), previous ablation times (de novo). LADs were measured in anterior-posterior (AP), right-left (RL), and superior-inferior (SI) directions from raw CT scans^13,14^.

### Time-To-Recurrence Days and Censoring

Patients were routinely followed up post-ablation to survey for AF recurrence. Event monitors or pacemaker interrogations were used for arrhythmia monitoring. The censoring limit in our analysis was set to 36 months after catheter ablation. Time-to-recurrence data and censor indicator were defined as follows:

- Patients with AF recurrence: time-to-recurrence was calculated as the time interval between the ablation date and the recurrence diagnosis date. Their censoring indicator was set to 1.
- Patient without recurrence: For patients with no documented recurrence within the 36-month follow-up period, time-to-recurrence was set to 36 months, and the censoring indicator was set to 0.
- Patients with lost to follow-up: For patients lost to follow-up before reaching the censoring limit, time-to-recurrence was defined as the time from ablation to the last available follow-up. Their censoring indicator was set to 0.

### Architecture of MARTA-AF

As shown in Figure 1, the network architecture of MARTA-AF consisted of an aggregated network architecture to integrate different input modalities^15^. MARTA-AF employed a ResNet architecture^16^ for the imaging branch and a fully-connected neural network (FNN) architecture for the tabular branch to analyze different data modalities. Encoded representations from both modalities were combined via a data fusion module to predict personalized risk parameters, denoted as µ_i_ and σ, from which individualized hazard function were derived. This semi-parametric approach contrasts with previous approaches, where the recurrence risk was directly predicted by AI^7,9^, enabling an efficient training in small-data settings. As shown in the right panel of Figure 1, estimated hazard functions were then used in a series of downstream analyses, including survival curves estimation, patient stratification at various time points, and interpretability analysis. More technical details are provided in Supplementary 1.

### Interpretability

To understand how MARTA-AF estimated the individualized risk, we applied a post-hoc attribution-based method to the tabular data branch. In that branch, Shapley values^17^ were computed to quantify the contribution of each input to the predicted risk. A positive Shapley value indicates a contribution to recurrence risk, whereas negative values reflect a protective effect. Tabular data were ranked based on their magnitude of impact.

### Statistical Analysis and Censoring

For patient characteristics, analyses were performed using the Python SciPy package. Normally distributed numeric data were presented in mean and standard deviation (SD) and non-normally distributed numeric data were reported as medians and interquartile ranges (IQR). Categorical data were presented in count and percentage. Group comparisons for continuous data were performed using the Kruskal–Wallis test or Mann–Whitney U test, as appropriate, while categorical data were compared using the χ^2^ test. Group comparisons for survival analysis were conducted using the log-rank test in the Python *lifeline* package.

Numeric data were divided into two groups by a threshold, followed by a log-rank test for hypothesis testing. The significance level was set as 0.05.

Model performance was evaluated using the mean and SD of the concordance index (C-index)^18^ and integrated Brier score (IBS) calculated from 5-fold cross-validation. In the AI interpretability section, the overall impact of a tabular input was quantified by the mean of its absolute Shapley values. The correlation of a tabular input and its Shapley value was quantified by Pearson’s correlation coefficient, r.

## Results

### Patient Characteristics

Baseline characteristics of the cohort are presented in Table 1. To understand the underlying data distribution, hypothesis tests were performed in patients with recurrence and no recurrence at 1 year after ablation for consistency with previous studies. Of these, 32 patients were excluded from the statistical analysis due to lost to follow-up at 12 months. In particular, these patients were only excluded in statistical analyses, while retaining them in the AI training and validation cohorts. Demographic analysis revealed that patients with AF recurrence were significantly taller (p-value=0.023, *), exhibited higher BMI (p-value=0.008, **) and greater body weight (p-value=0.001, **), compared to those without recurrence^19^. In addition, patients with OSA (p-value=0.027, *), PsAF (p-value=0.008, **), and enlarged LA were strongly associated with a higher chance of experiencing recurrence (p-value<0.001, ***).

**Table 1:** Patient characteristics of the cohort. 32 patients were excluded as they were lost to follow-up within a year. Normally distributed data are presented as mean with standard deviation (SD), and non-normally distributed data are presented as medians with interquartile range (IQR). Categorical data are presented as counts with percentages. *: p-value<0.05, **: p-value<0.01, ***: p-value<0.001. BMI: body mass index; CAD: coronary artery disease; OSA: obstructive sleep apnea, RF: radiofrequency; LAD: left atrial diameter; AP: Anterior-Posterior; RL: Right-Left; SI.: Superior-Inferior.

| Tabular Data | Recurrence (n=146) | No Recurrence (n=259) | p-value | Log-rank p-value |
| --- | --- | --- | --- | --- |
| **Demographics** |  |  |  |  |
| Age (years) | 66.05±9.06 | 64.75±10.90 | 0.293 | 0.368 (over 65) |
| Male | 100 (69%) | 164 (64%) | 0.329 | 0.318 |
| Height (cm) | 177.11±10.47 | 174.53±10.89 | 0.023, * | 0.009 (over 175), ** |
| Weight (kg) | 101.31±23.96 | 93.43±22.89 | 0.001, ** | 0.015 (over 85), * |
| BMI | 32.20±6.77 | 30.53±6.51 | 0.008, ** | 0.033 (over 27), * |
| **Comorbidities** |  |  |  |  |
| Stroke | 11 (8%) | 23 (9%) | 0.712 | 0.595 |
| CAD | 26 (18%) | 54 (21%) | 0.517 | 0.903 |
| OSA | 58 (40%) | 74 (29%) | 0.027, * | 0.073 |
| Hypertension | 102 (70%) | 164 (64%) | 0.263 | 0.156 |
| Diabetes | 27 (19%) | 47 (19%) | 1.000 | 0.724 |
| **AF-related covariates** |  |  |  |  |
| PsAF | 69 (48%) | 87 (34%) | 0.008, ** | 0.003, ** |
| AF Duration (years) | 3.0, [1.0, 7.0] | 3.0, [1.0, 7.0] | 0.826 | 0.219 (over 1) |
| **Procedural characteristics** |  |  |  |  |
| PVI | 98 (68%) | 174 (68%) | 1.000 | 0.385 |
| RF Ablation | 85 (59%) | 151 (59%) | 1.000 | 0.430 |
| de novo | 131 (89%) | 227 (87%) | 0.629 | 0.532 |
| **LADs** |  |  |  |  |
| LA-AP (mm) | 49.51±8.16 | 44.30±7.25 | <0.001, *** | <0.001 (over 51), *** |
| LA-RL (mm) | 79.67±8.72 | 74.83±9.08 | <0.001, *** | <0.001 (over 75), *** |
| LA-SI (mm) | 76.11±7.09 | 72.90±7.65 | <0.001, *** | 0.008 (over 75), ** |
| **Other covariates** |  |  |  |  |
| Pre-procedural LVEF | 55.98±10.91 | 56.89±10.04 | 0.522 | 0.408 (over 50) |
| CHA<sub>2</sub>DS<sub>2</sub> – VASc | 2.0, [2.0, 3.0] | 2.0, [1.0, 4.0] | 0.903 | 0.387 (over 1) |
| NT-proBNP (pg/mL) | 873.5, [295.25, 1930.0] | 473.0, [189.0, 1332.5] | 0.067 | 0.018 (over 600), * |

Log-rank tests were subsequently conducted to examine differences in time-to-recurrence in groups stratified by these covariates. Numeric data were divided by predefined thresholds shown in parentheses. The following factors were associated with a significantly reduced time-to-recurrence: height over 175 cm (p-value=0.009, **), weight over 85 kg (p-value=0.015, *), BMI over 27 (p-value=0.033, *), PsAF (p-value=0.003, **), received RF ablation (p-value=0.003, **), NT-proBNP over 600 (p-value=0.018, *), and enlarged LA (p-values<0.001, ***, in AP and RL direction and p-value=0.008, **, in SI direction).

### Examining MARTA-AF Performance in predicting the “*When*”

As described in the Introduction, predicting *when* an AF patient will experience recurrence post-ablation is clinically actionable. Predictive performance of a survival model is commonly evaluated using two complementary metrics: the C-index, which assesses how well a model ranks patients according to their relative risk, and the IBS, which measures the accuracy of predicted event probabilities over time. This section compares first the C-index and IBS of MARTA-AF with those of well-established clinical scores used as benchmarks, followed by evaluations of patient stratification based on their predicted risk.

Figure 2 presents the performance of MARTA-AF compared to benchmarks. Previous studies have indicated that *CHA*_2_*DS*_2_ − *VASc* and CAAP-AF are predictive of AF recurrence in different populations, with calculation methods detailed in ^20,21^; MARTA-AF was benchmarked against these clinical indices. In addition, the Cox proportional hazard model, the most widely used model in survival analysis, was adopted as another benchmark to predict time-varying recurrence risk. As shown in Figure 2a, the C-indices of *CHA*_2_*DS*_2_ − *VASc*, CAAP-AF, and the Cox model were 0.509, 0.608, and 0.608±0.050, respectively. Figure 2b shows IBSs of 0.259, 0.250, and 0.207±0.011 for the three benchmarks. Since the clinical scores do not require model training, SD is not applicable and thus not reported. The results suggest that these benchmark models exhibited low to modest performance, with some approaches marginally outperforming random guess (dashed line).

**Figure 2:**
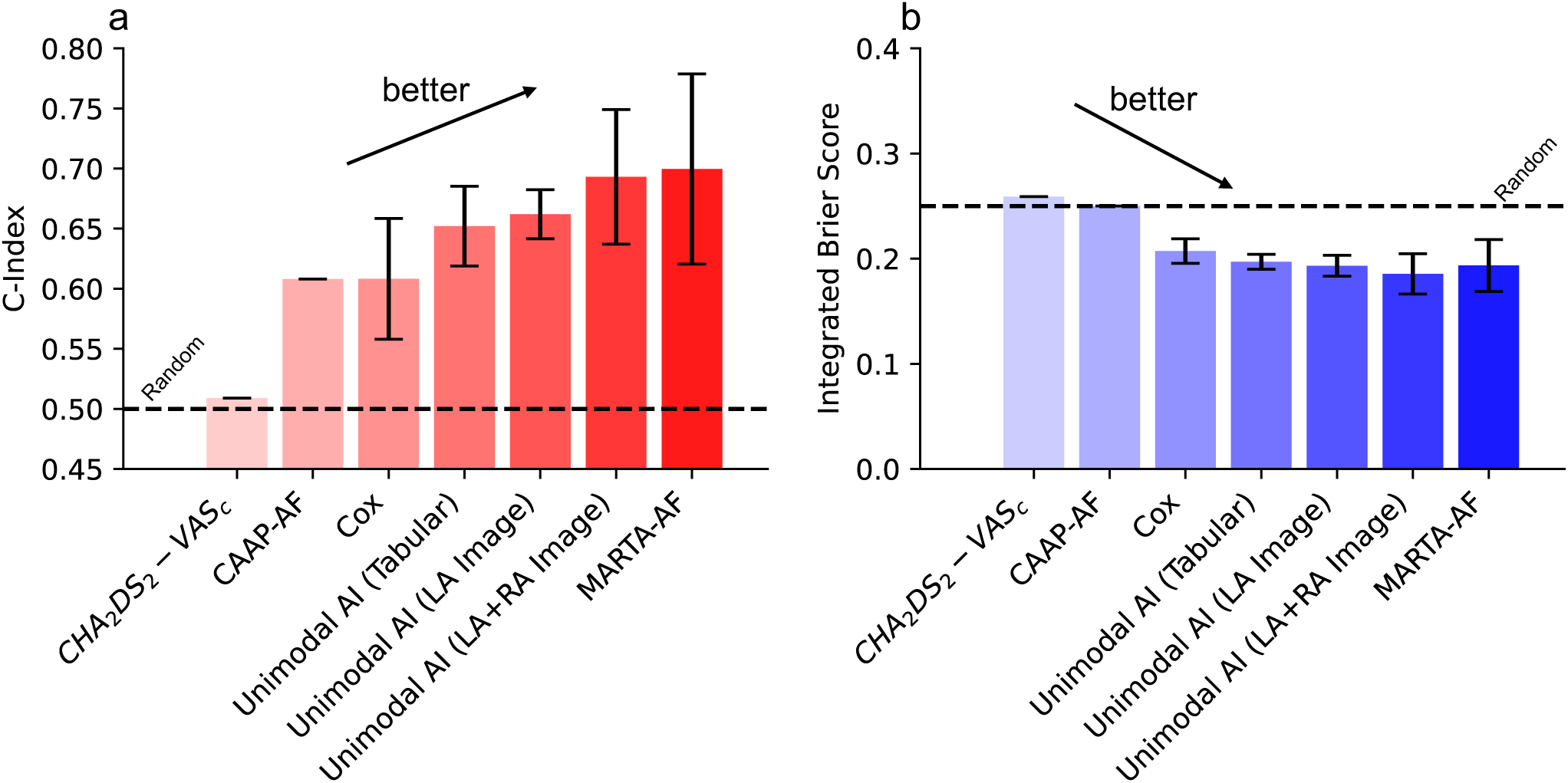
Performance of MARTA-AF (multimodal) compared against clinical scores, Cox model, and unimodal AI. a) C-index and b) integrated Brier score are presented for a series of benchmarks, including *CHA*_2_*DS*_2_*VASc* score, CAAP-AF, Cox proportional model, unimodal AI (tabular data, LA image, and LA+RA image) compared with MARTA-AF. Dashed lines indicated the performance of random guess.

The AI performance was then evaluated. We first compared the MARTA-AF performance with that of unimodal AI, where the imaging and tabular data branches were trained independently; this allowed us to assess the contribution of multimodal data inputs. For each unimodal AI, a linear layer with 2 neurons was appended to predict the personalized risk parameters µ and σ. The unimodal AI achieved C-indices of 0.652±0.033 for tabular data alone and 0.662±0.020 for LA images alone. Unimodal IBS scores were 0.193±0.009 and 0.198±0.010, accordingly. Notably, compared to the unimodal AI using only LA images, unimodal AI incorporating bi-atrial (LA+RA) images achieved a higher C-index of 0.693±0.056 and a lower IBS of 0.185±0.019. Finally, MARTA-AF that integrates LA and RA images and tabular data achieved the best performance, with a C-index of 0.703±0.079 and IBS of 0.190±0.024. These results highlight the benefits of developing an AI strategy that leverages multimodal medical data, and indicate that the RA structural information provides unique predictive value for AF time-to-recurrence. More details of AI discriminative performance over time are provided in Supplementary Figure 1.

Next, we evaluated the effectiveness of AI-based patient stratification by examining the survival curves in different risk groups. As a benchmark, Kaplan-Meier curves stratified by AF type are presented in Figure 3a. A significant difference in AF time-to-recurrence was observed between these groups (log-rank p-value=0.003, **). The restricted mean survival time (RMST) over the 3-year follow-up was 2.04 years for patients with PaAF and 1.68 years for PsAF patients, leading to a 0.36-year gap in population-wise recurrence-free time.

**Figure 3:**
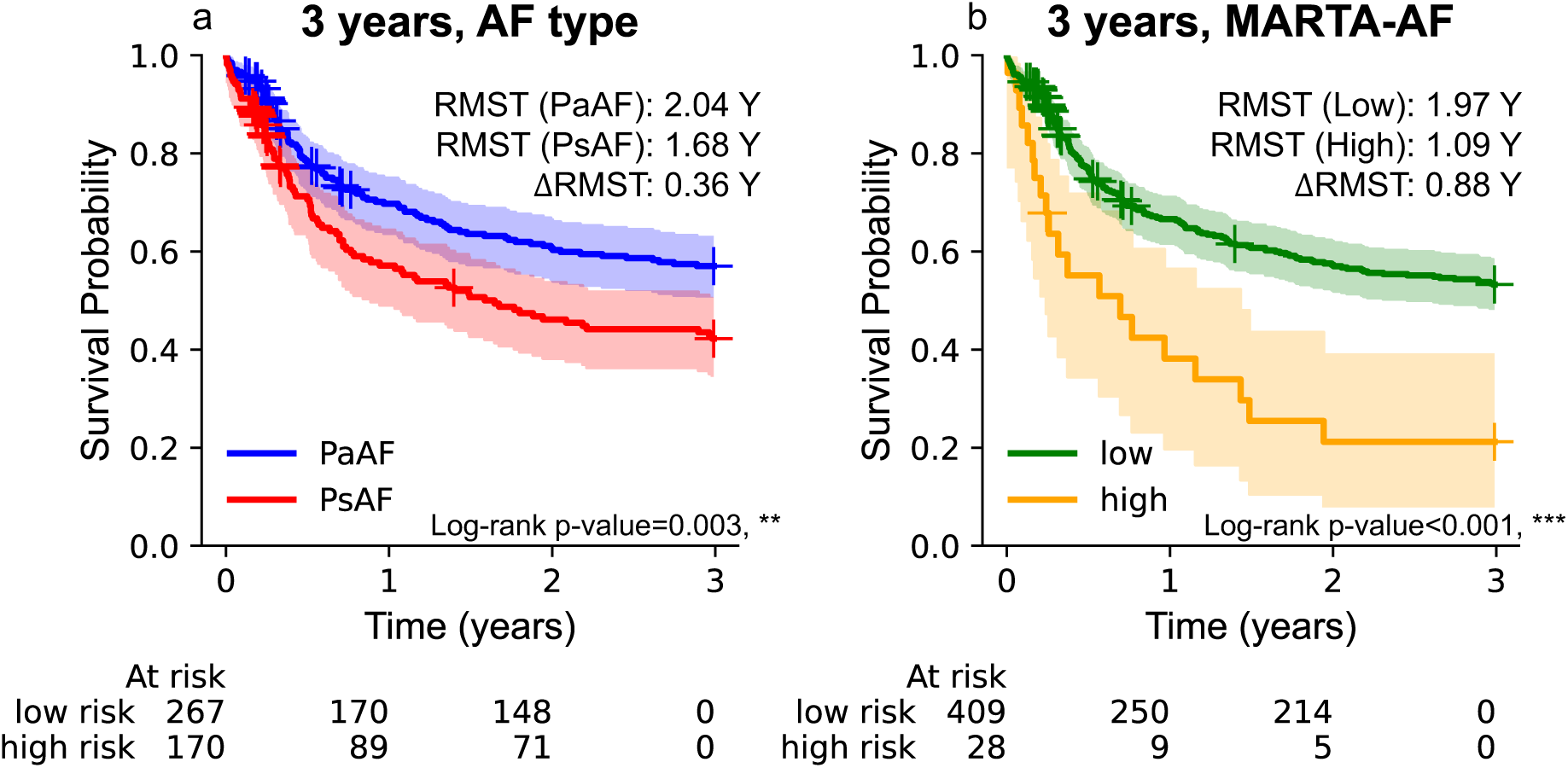
Kaplan-Meier plots with population-wise restricted mean survival time (RMST). Kaplan-Meier plots for patient stratification by a) AF type and b) risk estimated by the MARTA-AF. Patients in b) were stratified based on their risk at 3-year post-ablation. The number of low– and high-risk patients from 0-36 months is presented at the bottom. The log-rank test is performed to compare survival curves between risk groups. The cross marks represent censored data points.

In contrast, patient stratification based on MARTA-AF is presented in Figure 3b. Patients in the highest 25% percentile of cumulated risk at 3 years were classified as high risk, while the remaining patients were considered low risk. To differentiate the stratification by AF type, the Kaplan-Meier curves of MARTA-AF-stratified low and high-risk groups were plotted in green and yellow. This new stratification yielded a remarkably larger separation between risk groups: the RMST of the high-risk group significantly decreased from 1.68 years to 1.09 years, while the RMST in the low-risk group (RMST 1.97 years) was similar to the PaAF group. The survival probability of the high-risk group also dropped to around 20% in 3 years, in contrast to above 45% in the PsAF group. The new stratification resulted in a larger difference of 0.88-year in survival time and a stronger statistical significance (log-rank p-value<0.001, ***) between these groups. Taken together, these results indicated that MARTA-AF enables a more effective and clinically accurate stratification than AF type.

To gain insights into the characteristics of the high– and low-risk groups, a group-wise comparison is presented in Table 2. As expected, the proportion of patients experiencing recurrence in the high-risk group (68%) was significantly higher than that of the low-risk group (28%) (p-value<0.001, ***). The distribution of the baseline characteristics is largely consistent with that in Table 1. Notably, several tabular inputs – including sex, hypertension, RF ablation, history of previous ablation (de novo), and NT-proBNP – exhibited significant differences between the AI-based stratification. The statistical significance in sex and hypertension may reflect the underlying selection or demographic biases. Regarding RF ablation, the association could be attributed to the clinical practice routine that RF ablation is often performed in patients with enlarged left atria, a population inherently at higher risk of recurrence^22^.

**Table 2:** Characteristics of high and low risk patients estimated by MARTA-AF.

| **Tabular Data** | High risk (n=97) | Low risk (n=308) | p-value |
| --- | --- | --- | --- |
| **Demographics** |  |  |  |
| Age | 67.10±8.76 | 64.62±10.66 | 0.064 |
| Male | 77 (80%) | 187 (61%) | 0.001, ** |
| Height (cm) | 180.05±9.41 | 174.02±10.82 | <0.001, *** |
| Weight (kg) | 108.84±22.37 | 92.31±22.55 | <0.001, *** |
| BMI | 33.62±7.02 | 30.35±6.33 | <0.001, *** |
| **Comorbidities** |  |  |  |
| Stroke | 5 (6%) | 29 (10%) | 0.214 |
| CAD | 19 (20%) | 61 (20%) | 1.000 |
| OSA | 45 (47%) | 87 (29%) | 0.001, ** |
| Hypertension | 76 (79%) | 190 (62%) | 0.002, ** |
| Diabetes | 22 (23%) | 52 (17%) | 0.228 |
| **AF-related covariates** |  |  |  |
| PsAF | 66 (69%) | 90 (30%) | <0.001, *** |
| AF Duration (years) | 4.0, [1.0, 8.0] | 2.5, [1.0, 7.0] | 0.141 |
| **Procedural characteristics** |  |  |  |
| PVI | 59 (61%) | 213 (70%) | 0.138 |
| RF Ablation | 67 (70%) | 169 (55%) | 0.013, * |
| de novo | 80 (82%) | 278 (90%) | 0.045, * |
| **LADs** |  |  |  |
| LA-AP (mm) | 53.11±6.87 | 43.99±7.01 | <0.001, *** |
| LA-RL (mm) | 84.82±7.97 | 73.97±8.02 | <0.001, *** |
| LA-SI (mm) | 79.09±6.62 | 72.47±7.20 | <0.001, *** |
| **Other covariates** |  |  |  |
| Pre-procedural LVEF | 55.82±10.20 | 56.80±10.42 | 0.241 |
| CHA<sub>2</sub>DS<sub>2</sub>VASc | 3.0, [2.0, 4.0] | 2.0, [1.0, 4.0] | 0.104 |
| NT-proBNP | 903.0, [366.5, 1930.0] | 470.5, [188.25, 1424.5] | 0.024, * |
| Recurrence | 60 (62%) | 86 (28%) | <0.001, *** |

### MARTA-AF effectively stratifies patients for AF recurrence at various time points

Risk of AF recurrence manifests in a non-proportional fashion: some patients maintain a low risk during the entire follow-up period, whereas some may have low recurrence risk during the blanking period but become subject to a sudden increase in recurrence risk shortly thereafter. To illustrate how MARTA-AF captures these risk patterns, we showcase representative patient-level predictions. Figure 4 a-c Illustrates model predictions for three patients: a) one who remained recurrence-free 3 years post procedure, b) a patient with early recurrence (less than 3 months), or c) a patient with late recurrence (after 12 months of ablation).The blue and purple curves indicate the survival probability and hazard function predicted by MARTA-AF. For patients who experienced AF recurrence, the true recurrence time is indicated by a black solid line; for patients without AF recurrence, the censoring time (marked in red) was set at 36 months and the hazard predicted by MARTA-AF stayed low throughout follow-up, with survival probability above 70% at 3 years. In contrast, the patient with early recurrence (black line) exhibited a very high hazard shortly after ablation, consistent with the observed recurrence event. For the patient with late AF recurrence, the event occurred at around 15 months. The predicted hazard increased rapidly from 0, peaked at approximately 1 year, and gradually decrease over year 2 to 3. This indicates that MARTA-AF is able to capture this non-proportionality of the risk over time, with high-risk periods clearly delineated.

**Figure 4:**
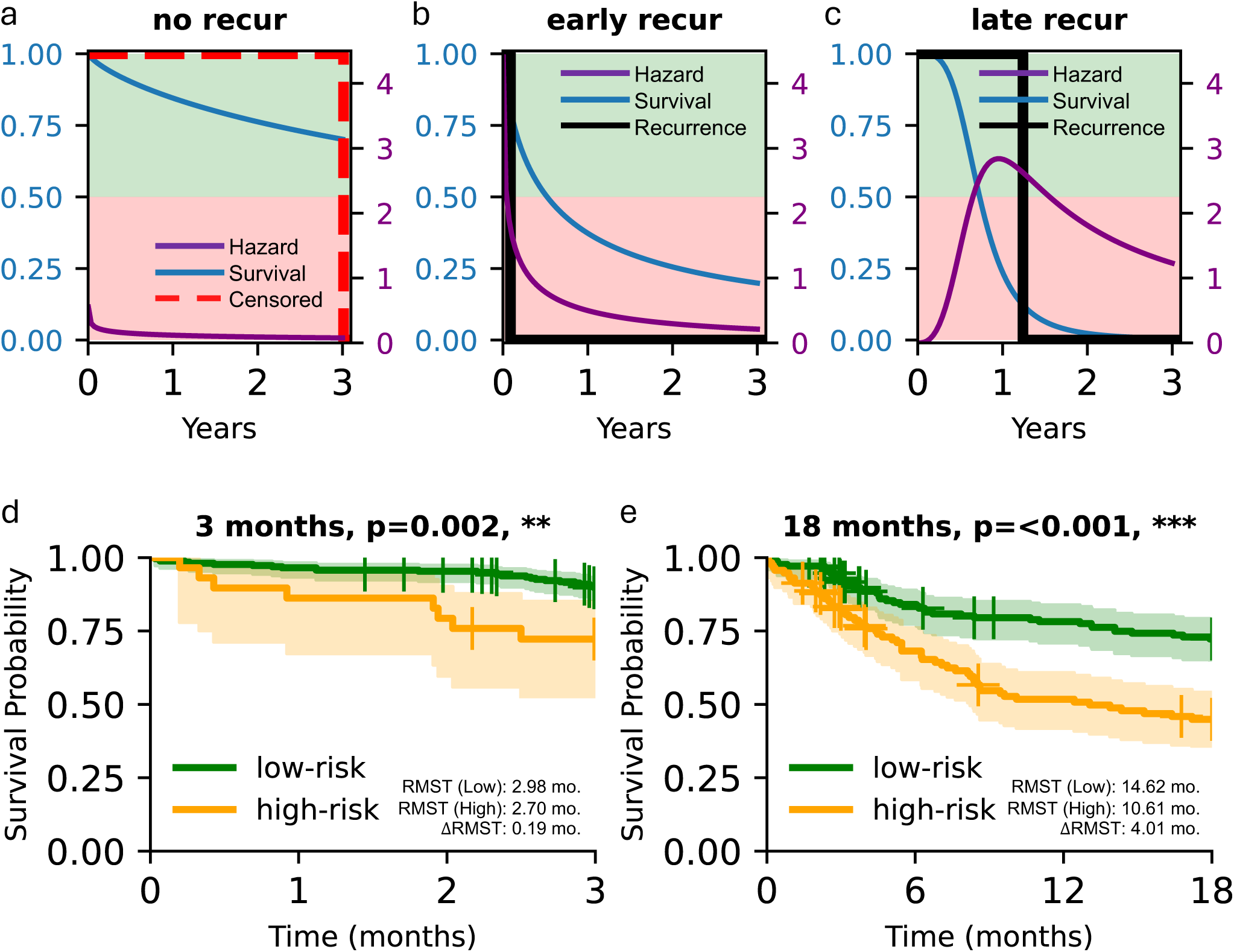
Individual risk estimates and patient stratification at 3 months (early) and 18 months (late) post-ablation from MARTA-AF. Three cases with hazard and survival prediction for a) no-recurrence, b) early-recurrence, and c) late-recurrence. Kaplan-Meier curves based on AF recurrence risk prediction at d) 3 months and e) 18 months post-ablation.

In light of the non-proportionality in AF recurrence risk, the effectiveness of patient stratification should be examined at multiple time points. Herein, we chose to estimate risk at 3 and 18 months to represent its performance (Figure 4 d and e). Following the same stratification strategy, patients were stratified into high and low risk groups. As shown in Figure 4d, the 3-month survival probability of the low-risk group (green, RMST=2.98 month within 3 months) exceeded 90%, whereas the high-risk group (yellow, RMST=2.70 month within 3 months) exhibited a significantly lower (p=0.002, **) probability of approximately 75%. A great separation was observed for survival curves at 18 months in Figure 4e (log-rank p-value<0.001, ***): the low-risk group maintained a higher survival probability of around 76% (RMST=14.62 months), while the high-risk group decreased to less than 50% (RMST=10.61 months).

### Predicting the *When* in subpopulations

It is expected that a model should perform consistently across different populations to minimize bias and ensure fairness. MARTA-AF Performance in different populations is presented in Figure 5. MARTA-AF predictive performance was compared with *CHA*_2_*DS*_2_ − *VASc* and CAAP-AF across subgroups in age, sex, AF type, and time-to-AF-recurrence. For reference, the mean and SD of the MARTA-AF performance in the overall cohort are indicated in Figure 5 by the dashed line and shaded in light blue. As shown in Figure 5a, the C-indices in all subpopulations were larger than the benchmarks, indicating that MARTA-AF has a better ability at ranking AF recurrence risk than clinical scores. Additionally, predictive performance of MARTA-AF remained largely consistent with its overall cohort performance across most subgroups, with modest reduction in C-index in the group with age < 65.

**Figure 5:**
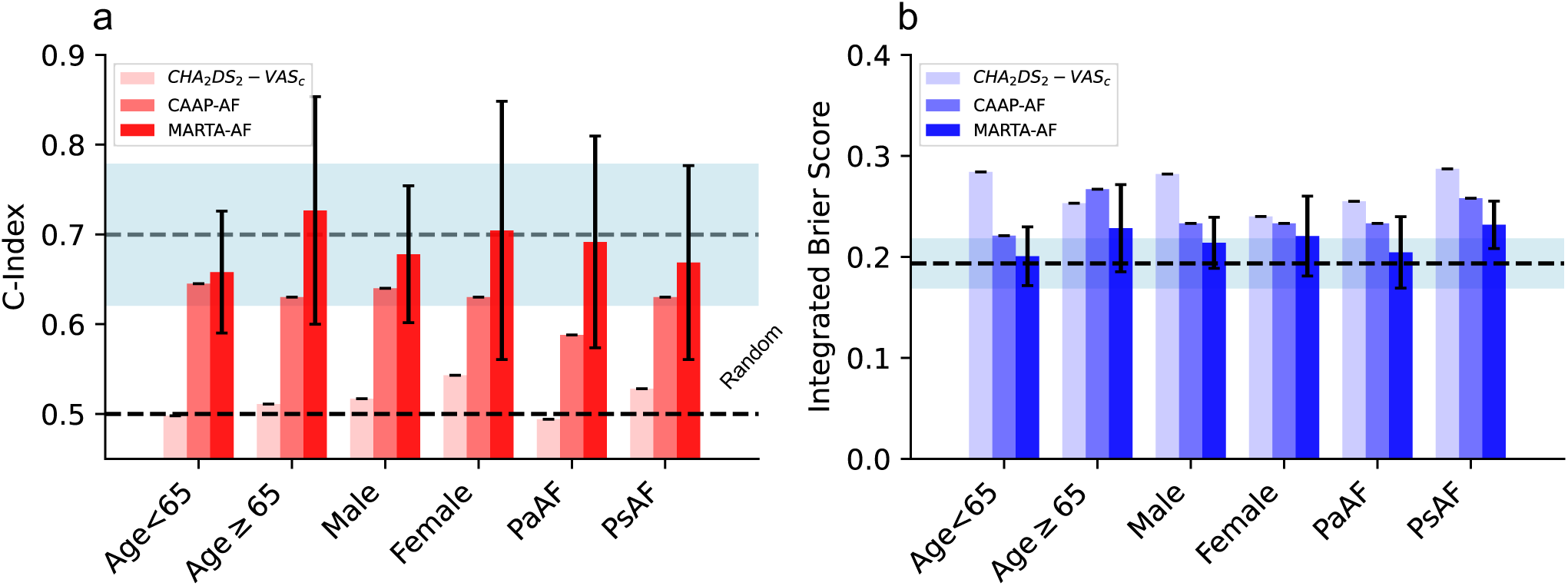
Performance of benchmarks and MARTA-AF in subpopulations. a) C-index and b) IBS in populations stratified by age, sex, and AF type. C-index and integrated Brier score are presented as mean and SD. The dashed line and blue band indicate the mean and SD of MARTA-AF performance in the overall cohort.

IBS scores in Figure 5b demonstrated that of MARTA-AF were the lowest among the subgroups in age, sex and AF type. Additionally, the SDs in subgroup performance metrics were noticeably larger than that in the overall cohort, suggesting an increased variability likely due to the limited size in these groups.

Next, we examined the effectiveness of the MARTA-AF patient stratification in these subpopulations (Figure 6). All the survival curves exhibited statistical significance (p-value<0.05) by log-rank tests. The low-risk groups (green curves) exhibited a 3-year survival probability over 60%, whereas the high-risk groups (orange curves) were far below 40%, except for the male group. The minimum difference in RMST was observed in the male group (0.46 year) and the largest survival separation was in the group with Age<65 (ΔRMST=0.8 year in 3-year follow-up). Overall, these results suggested that MARTA-AF is fair across most clinically relevant subgroups and could effectively stratify AF time-varying recurrence risk in diverse populations.

**Figure 6:**
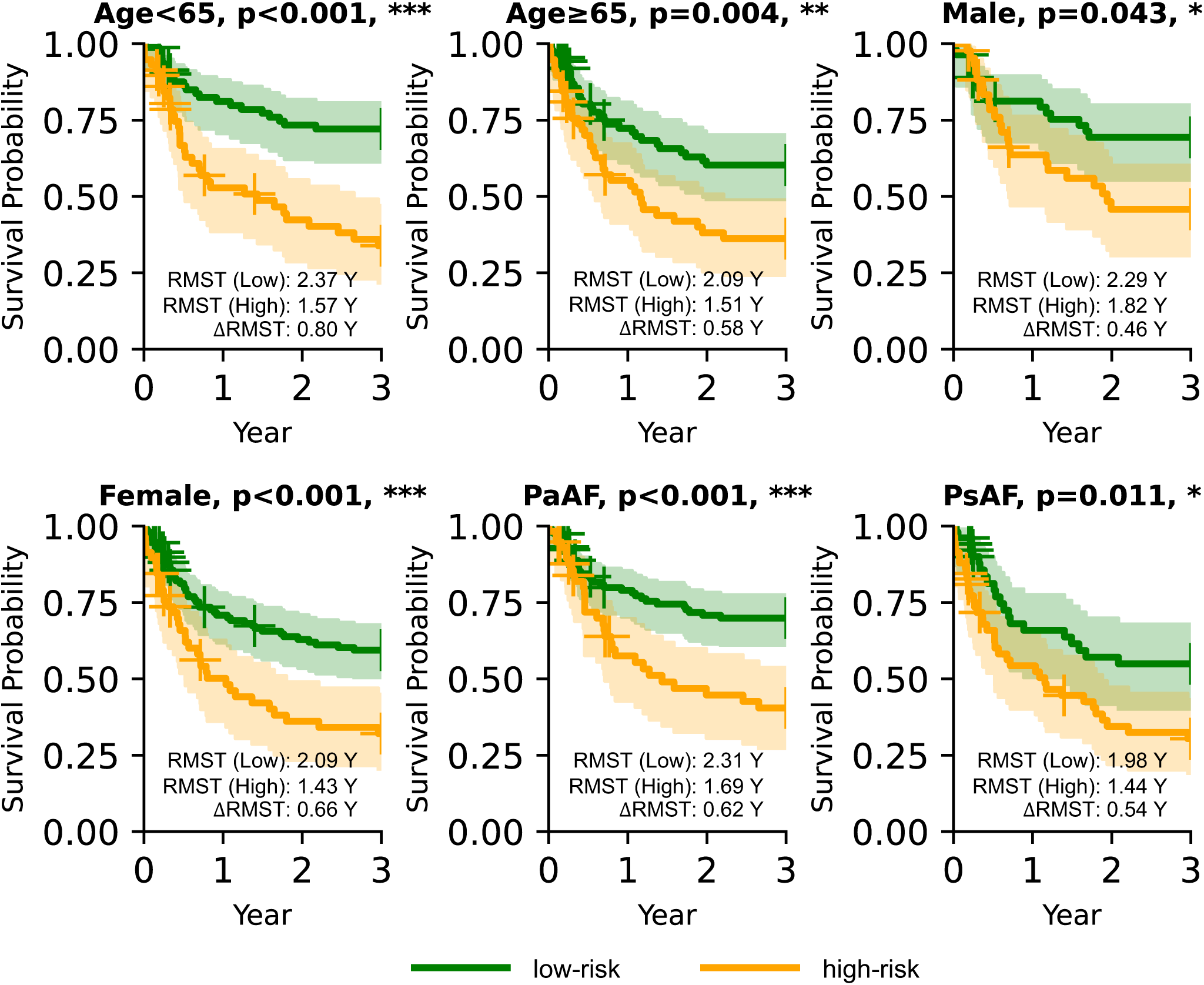
MARTA-AF stratified risk in subpopulations. Kaplan-Meier curves based on MARTA-AF-predicted risk for subpopulations.

**Figure 6:**
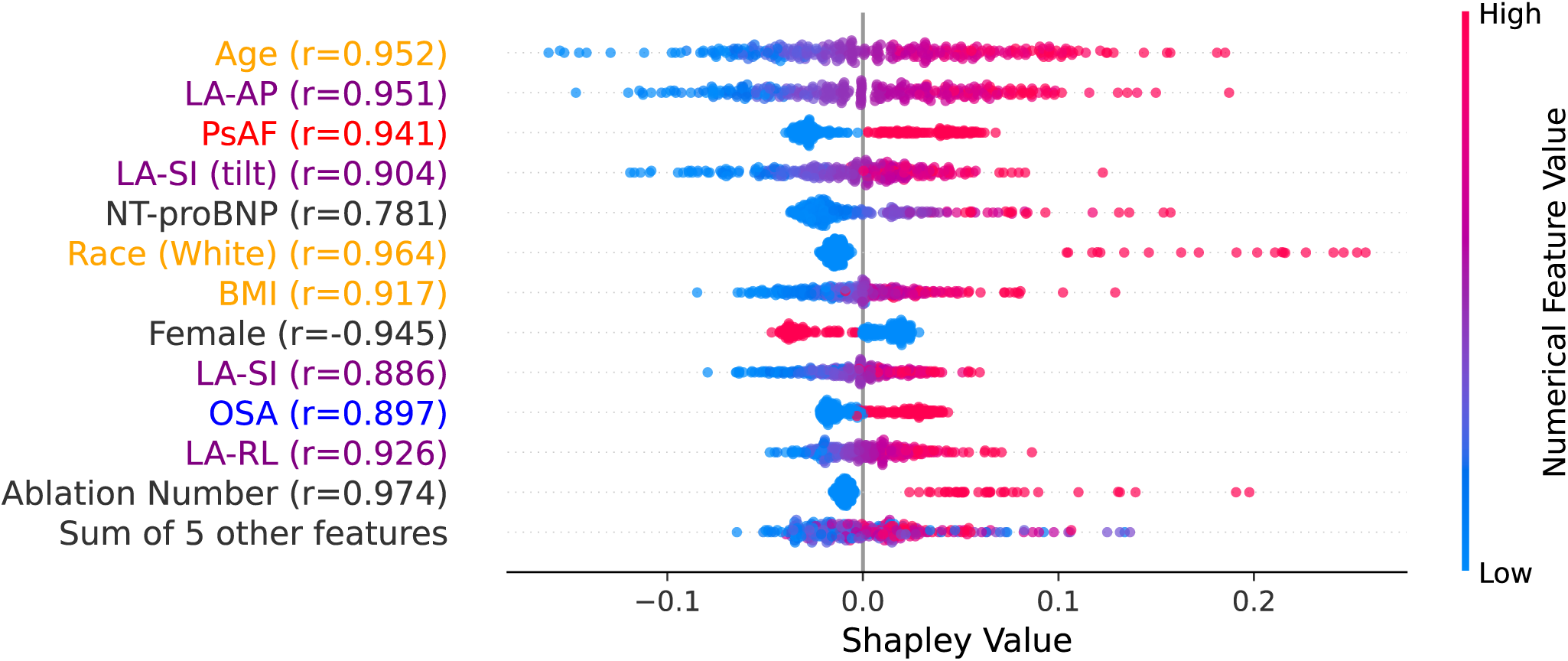
Tabular Inputs Interpretation. Shapley values of the tabular data are presented with a beeswarm plot. We present the top 12 most impactful tabular inputs (quantified by the mean of absolute Shapley values) along with their Pearson’s correlation coefficient (r) between the Shapley values and recurrence risk. The data are classified as either demographic (orange), comorbidities (blue), AF-related (red), LADs (purple), or others (black).

### Interpretability Quantifies the Impact of Tabular Data on Survival Probability

Interpreting AI algorithms is crucial for building trustworthy models and facilitating their deployment in clinical practice. To dive deeper into how MARTA-AF predicted recurrence risk, we examined the tabular branch using a Shapley-based attribution approach. The top 12 tabular inputs, ranked by their mean Shapley values, are presented in Figure 7, along with their Pearson’s correlation r with cumulative recurrence risk. Among these tabular inputs, age (r=0.952), LA-AP (r=0.951), PsAF (r=0.941), and LA-SI (r=0.904) were the most important factors to the increased recurrence risk.

**Figure 7:**
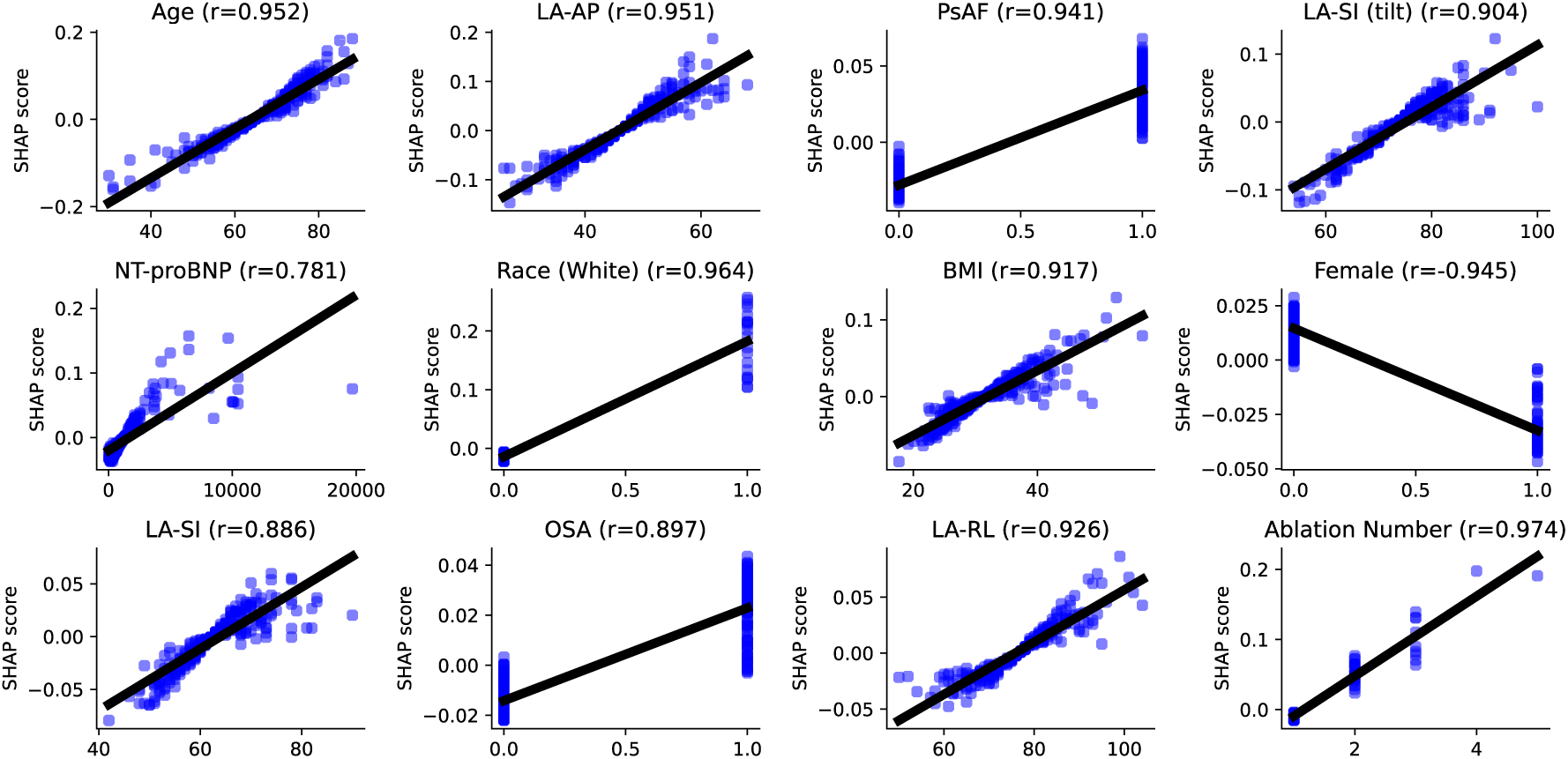
Shapley values for the top 12 most influential tabular inputs and linear fitting.

Detailed association between Shapley values (scores) and the tabular data value for the 12 most influential features are shown in Figure 8. Linear regression was deployed to characterize the relationships. Most tabular data exhibited a positive correlation, indicating that higher feature values were generally associated with increase in AF recurrence risk. Notably, for numeric data, a large variation in Shapley value was observed at high values of LA size, BMI, and NT-pro-BNP. This finding suggests that although atrial enlargement and elevated value in risk factors are statistically associated with a higher recurrence risk, their contribution to recurrence risk may vary for different individuals, as arrhythmia was successfully controlled by catheter ablation in many patients in our cohort. Overall, these results were consistent with data in Table 1 and results from previous studies.

## Discussion

This study introduces MARTA-AF, a novel framework integrating multimodal AI with survival analysis to predict, at the individual patient level, the timing of AF recurrence following catheter ablation. The principal findings are fourfold: (1) MARTA-AF successfully predicted time-varying recurrence risk up to three-years post-ablation; (2) MARTA-AF demonstrated its fairness and consistent effectiveness across clinically relevant subgroups, including sex, age, and AF type; (3) incorporation of RA shape features improved MAARTA_AF performance; and (4) analyses of patient characteristics across risk strata, combined with AI interpretability methods, identified a set of recurrence predictors well-aligned with those reported in prior independent studies.

By delivering individualized, pre-procedural time-to-recurrence risk forecasts, MARTA-AF has the potential to transform post-ablation AF management from a reactive, one-size-fits-all approach to a proactive, personalized paradigm. Such forecasts could meaningfully inform clinical decision-making across several domains — including optimization of anticoagulation strategies, timing of repeat ablation procedures, and the design of remote rhythm monitoring and follow-up protocols tailored to each patient’s risk trajectory. Critically, because risk is assessed prior to the ablation procedure itself, clinicians are equipped to individualize management strategies before committing to an intervention. For patients classified as low-risk, catheter ablation can be pursued with greater confidence. Conversely, for patients with a history of multiple prior ablations who are identified as likely non-responders — and in whom high hazard is predicted early in the post-ablation period — a more conservative strategy should be considered, including deferral of further ablation in favor of alternative rhythm control approaches.

AF recurrence can manifest anywhere from a days^23^ to several years^24^ following ablation. While time-varying risk forecasts would offer substantially greater clinical utility than single time-point predictions, existing prognostic AI models have focused predominantly on the binary question of whether AF will recur rather than when. Initial attempts at time-to-recurrence prediction include Zhou et al^25^, who developed an AI-based survival model using four clinical covariates, and Schroder et al^26^, who proposed an AI approach to predict AF recurrence-related hospitalization risk from clinical data alone. Despite these early efforts, a critical gap remains: no prior model has leveraged multimodal data — integrating both medical imaging and clinical covariates — for comprehensive, individualized time-to-recurrence analysis. MARTA-AF directly addresses this unmet need by combining bi-atrial imaging with clinical inputs within a unified multimodal AI and survival analysis framework, enabling personalized recurrence risk forecasting across a clinically meaningful time horizon.

Prior AF recurrence prediction has focused predominantly on LA morphology, despite growing evidence that the RA is mechanistically implicated in AF and its recurrence^27^. Structurally, RA remodeling — including enlargement^11^ and increased sphericity^28^ — has been associated with elevated recurrence risk, likely mediated through enhanced fibrosis^28^ and abnormal conduction velocity and block^29^. These findings suggest that RA morphology carries recurrence-relevant information that LA-only models may fail to capture. Nevertheless, very few predictive frameworks have explicitly incorporated RA shape features^30,31^. The present study addresses this gap directly: our results demonstrate that combining LA and RA morphology significantly improves predictive performance over LA-only models, confirming that RA shape contributes independent and clinically meaningful information to AF recurrence risk prediction.

### limitations

Our study has several limitations. Although MARTA-AF achieved a favorable C-index, IBS is relatively modest. This shows that the model is stronger at ranking patients by their recurrence risk than estimating recurrence time. In addition, MARTA-AF was developed based on pre-procedural data, whereas information of post-ablation managements, which could influence the risk, is not included in our dataset. These limitations likely contribute the inherent stochasticity and uncertainty of time-to-AF-recurrence, and incorporating additional relevant factors may help improve model performance. Furthermore, although we have incorporated LA and RA morphology, fibrosis may be beneficial to include as the fibrosis distribution is mechanistically associated with recurrence reentrant drivers^32^. Incorporating fibrotic features reflected in late-gadolinium enhanced (LGE) MRI^30^ could provide additional AF recurrence-related insights that could be valuable for risk prediction. In addition, medical data, such as 12-lead ECG signals^16^ or intracardiac electrogram^9^ and have been shown to be predictive of AF recurrence as they contain information that reflects the underlying electrophysiological function. MARTA-AF is readily capable of learning from such data thanks to its modular design: new imaging modalities or time-series signals can be integrated via dedicated branches, while additional clinical covariates can be incorporated into the existing tabular branch. Finally, external validation of MARTA-AF has not yet been performed, due to the scarcity of cohorts with pre-procedural bi-atrial imaging data of the type used in this study. Thus, external validation in prospective cohorts will be the important next step.

## Data Availability

Source data can be provided by request to the corresponding authors and potentially after an IRB for sharing data is approved.

## Ethics

MARTA-AF was developed based on the Johns Hopkins Atrial Fibrillation Registry. JHIRB approved this study because the study uses retrospective and unidentified data.

## Author Contributions Statement

M.Y., H.C., D.S., and N.T. designed research;

R.Y., J.M., L.T., C.O., S.S., C.C., and C.W. performed data curation;

M.Y., C.P., Z.A., S.C., A.L., J.R., A.Y., J.N., and E.K. performed medical images preprocessing;

M.Y. and C.L. performed network training;

M.Y., M.M., H.C., D.S., and N.T. analyzed data; All authors contributed to writing the paper.

## Supplementary

### 1. Multimodal AI and Survival Analysis

We employed modality-specific network architectures to learn from each input modality. The imaging branch consisted of a 4-layer 3D ResNet^16^ to analyze LA morphology from imaging data. The tabular branch employed a feedforward neural network (FNN, 3 layers, 32 neurons in each layer). Categorical data were one-hot encoded, and numeric data were normalized by subtracting the mean and dividing the SD.

A key ingredient of a survival analysis was the estimation of its survival function (S), which characterizes the cumulative survival probability of a patient over time. Its complement, F = 1 − S, is the cumulative distribution function of risk that describes the cumulative risk of recurrence at a time point. A special non-proportional risk model was chosen from Popescu et al.^18^. Unlike traditional proportional hazard models, which assume that the relative risk between patients remains constant over time, the non-proportional model allows for time-varying risk rankings. For example, patient A may have a higher recurrence risk than patient B at 3 months, but B could be at a higher risk than A at 12 months. This kind of risk reordering cannot be captured by proportional hazard models but is accommodated by the non-proportional model. The survival function in this framework was defined as follows:

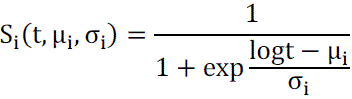

where t is the time at which the recurrence risk is to be estimated. Individualized survival prediction for an AF patient i was achieved by parameterizing the survival function with patient-specific values of µ_i_ and σ_i_. In our model, these parameters were predicted by a FNN data fusion module, which integrated information from both the CT segmentation and tabular data. To estimate the recurrence risk at a specific time point t, one simply inputs the patient’s medical images and tabular data into the model and calculates the cumulative distribution function of risk (F) based on predicted µ and σ.

The network was trained to minimize the negative log-likelihood of this survival model:

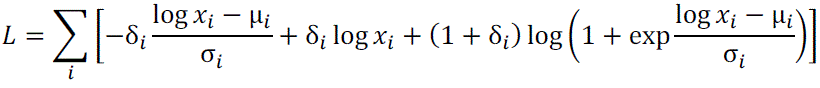

Where δ_i_ is the event indicator for patient i (with δ_i_ = 1 if recurrence is observed and δ_i_ = 0 if censored), and x_i_ is the time-to-recurrence from data. This formulation enables the model to handle right-censored data while learning individualized recurrence risk over time.

### 2. Predicting *Whether* at multiple time points

**Supplementary Figure 1:**
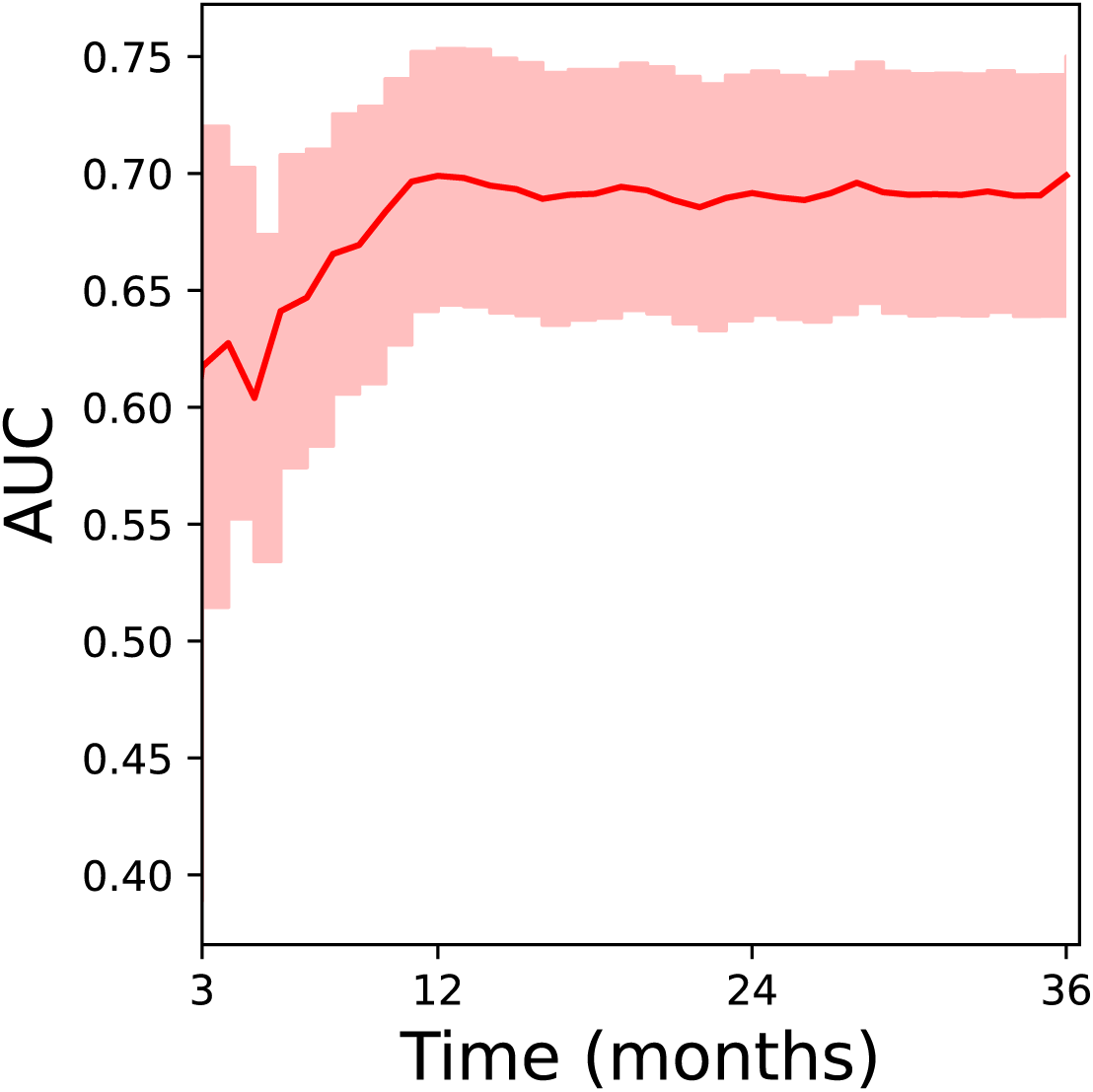
AUC score over time.

In this section, we examined the discriminative performance of the AI model over time by calculating the AUC score from 3 to 36 months after ablation. Note that AUC is equal to the C-index when assessed at a single time point. As shown in Supplementary Figure 1, the AUC score increases from 0.62 at 3 months to 0.70 at 11 months and remains stable around 0.70 from 12 to 36 months. The shaded area indicates the confidence interval of the AUC score. One possible explanation of the model’s relatively lower performance between 3 and 12 months is the impact of loss to follow-up and the clustering of censored data during this interval. This pattern of censoring effectively reduced the number of recurrence events, potentially introducing bias and increasing the variance in the estimation of the model and its performance.

Variations in performance for AF recurrence prediction have been observed across different studies. Performance of prediction AF recurrence in previous studies was often quantified by Area Under the Receiver Operating Characteristic (AUC), a standard statistical metric for evaluating classification ability. Razeghi et al.^7^ reported an AUC of 0.650 in a cohort of 321 AF patients using the *CHA*_2_*DS*_2_ − *VASc* score and Kornej et al.^33^ reported a slightly lower AUC of 0.572 for the *CHA*_2_*DS*_2_ − *VASc* score in a cohort of 879 AF patients. In contrast, in our cohort, the same *CHA*_2_*DS*_2_ − *VASc* score yielded a substantially lower AUC of 0.509, indicating that this risk score was not predictive of AF recurrence in our population. Similarly, performance also varied among AI-based approaches that learn from medical images and tabular data. Razeghi et al.^7^ reported an AUC of 0.82±0.14, whereas Kim et al.^6^ reported an AUC of 0.61 in a cohort of 527 AF patients. To provide a fair comparison, we implemented the architecture and training procedure described in Razeghi et al.^7^, yet observed a lower performance (AUC of 0.68) in our cohort. These discrepancies underscore the substantial impact of intra– and inter-cohort heterogeneity on model performance. Variations in patient characteristics, clinical settings, and data quality could significantly influence both traditional and AI-based risk prediction models. Therefore, a lower AUC value does not necessarily indicate inferior model performance in predicting AF recurrence.

